# No association between circulating levels of testosterone and sex hormone-binding globulin and risk of COVID-19 mortality in UK biobank

**DOI:** 10.1101/2020.09.11.20191783

**Authors:** Xikang Fan, Jing Yang, Jiayu Wang, Cheng Yin, Meng Zhu, Hongxia Ma, Guangfu Jin, Zhibin Hu, Hongbing Shen, Dong Hang

## Abstract

**Background:** Sex-disaggregated data suggest that men with coronavirus disease 2019 (COVID-19) are more likely to die than women. Whether circulating testosterone or sex hormone-binding globulin (SHBG) contributes to such sex differences remains unknown.

**Objective:** To evaluate the associations of circulating total testosterone (TT), free testosterone (FT), and SHBG with COVID-19 mortality.

**Design:** Prospective analysis.

**Setting:** UK Biobank.

**Participants:** We included 1306 COVID-19 patients (678 men and 628 women) who had serum TT and SHBG measurements and were free of cardiovascular disease or cancer at baseline (2006-2010).

**Main outcome measures:** The death cases of COVID-19 were identified from National Health Service death records updated at 31 July 2020. Unconditional logistic regression was performed to estimate the odds ratio (OR) and 95% confidence intervals (CI) for mortality.

**Results:** We documented 315 deaths of COVID-19 (194 men and 121 women). After adjusting for potential confounders, we did not find any statistically significant associations for TT (OR per 1-SD increase = 1.03, 95% CI: 0.85-1.25), FT (OR per 1-SD increase = 0.95, 95% CI: 0.77-1.17), or SHBG (OR per 1-SD increase = 1.09, 95% CI: 0.87-1.37) with COVID-19 mortality in men. Similar null results were observed in women (TT: OR per 1-SD increase = 1.10, 95% CI: 0.85-1.42; FT: OR per 1-SD increase = 1.10, 95% CI: 0.82-1.46; SHBG: OR per 1-SD increase = 1.16, 95% CI: 0.89-1.53).

**Conclusions:** Our findings do not support a significant role of circulating testosterone or SHBG in COVID-19 prognosis.

## Introduction

The coronavirus disease 2019 (COVID-19) pandemic is a global health emergency that has led to more than 870,000 deaths worldwide (https://covid19.who.int/). According to sex-disaggregated data from many affected countries, there seem to be sex differences in severity and mortality of the disease (1-3). Men with COVID-19 are more likely to need intensive care and to die than women, with an overall case-fatality ratio of approximately 1.7 (4). Several hypotheses have been proposed to account for the difference, such as smoking, co-morbid conditions, immune responses, and sex hormones (4, 5). As the most abundant circulating androgen in men, testosterone has been suggested as an immunosuppressive factor by inhibiting B lymphopoiesis, T-helper 1 differentiation, and inflammatory cytokine production (6, 7). However, epidemiologic data are sparse regarding the association between circulating testosterone levels and COVID-19 prognosis in either men or women.

Most circulating testosterone is tightly bound to sex hormone-binding globulin (SHBG) or weakly bound to albumin, and a minor amount exists as free testosterone (FT) (8). In the current study, we leveraged the UK Biobank resource, with recently released data on COVID-19 tests and updated death records, to examine the associations of total testosterone (TT), calculated FT, and SHBG with mortality among COVID-19 patients.

## Methods

### Study design and participants

UK Biobank enrolled over 500,000 community-dwelling individuals (aged 37-73 years) across the UK during 2006 to 2010 (9). At the baseline assessment, participants completed self-administered touchscreen questionnaires, had physical measurements by qualified nurses, and provided blood samples.

According to the Public Health England database linked to UK biobank updated at 3 August 2020, a total of 9773 participants had undergone severe acute respiratory syndrome coronavirus 2 (SARS-CoV-2) tests, among whom 1638 were diagnosed with COVID-19. We also included 161 participants who died from COVID-19 according to the National Health Service (NHS) death records. Exclusion criteria included COVID-19 patients who had no available data on serum TT (n=286) or SHBG (n=309), who had a self-reported history of cardiovascular disease (n=176) or cancer (n=138) at blood draw, and who died from other causes (n=22). Finally, 1306 patients with COVID-19 were included in the analysis, consisting of 678 men and 628 women.

### Assessment of biomarkers

Serum concentrations of TT and SHBG were measured by chemiluminescent immunoassays (Beckman Coulter DXI 800), with analytical ranges of 0.35-55.52 nmol/L and 0.33-242 nmol/L, respectively. A colorimetric method was used to assay albumin (Beckman Coulter AU5800), with an analytical range of 15-60 g/L. Overall, the average coefficients of variation for TT, SHBG, and albumin ranged from 3.66-8.34%, 5.22-5.67%, and 2.09-2.20%, respectively. Moreover, each assay was registered with an external quality assurance scheme to verify the accuracy, showing that more than 95% of participated distributions for these biomarkers were good or acceptable. FT was calculated using a validated algorithm based on TT, SHBG, albumin, and the association constants for the binding of testosterone to SHBG and albumin (10, 11)

### Ascertainment of COVID-19 Mortality

Date and cause of death were obtained through linkage to national death registries. Primary cause of mortality was defined using the 10th revision of the International Statistical Classification of Diseases, and the outcome of our analysis was mortality due to COVID-19 (U07). Available death data we used were last updated at 31 July 2020.

### Ascertainment of Covariates

Information on age, sex, education, ethnicity, smoking status, and alcohol consumption was self-reported in the baseline questionnaires. Townsend deprivation index was evaluated by incorporating measures of unemployment, household overcrowding, and non-ownership of home or car. Body mass index (BMI) was calculated as weight in kilograms divided by height in meters squared derived from physical measurements. Total physical activity was assessed by total metabolic equivalent task minutes per week for all activity, including walking, moderate, and vigorous activity.

### Statistical Analysis

Testosterone and SHBG concentrations were log-transformed to improve normality. Because the date of SARS-CoV-2 testing did not represent the date of getting infected, the survival time from infection to death or the last follow-up could not be accurately calculated for each patient, rendering Cox regression models inapplicable in the current analysis. We used unconditional logistic regression models to estimate odds ratios (OR) and 95% confidence intervals (CI), according to quintiles and continuously per 1-standard deviation (SD) increment in hormone concentrations. The model was first adjusted for major covariates including age at infection, sex, and ethnicity (simple model), then further adjusted for Townsend deprivation index, BMI, smoking status, alcohol drinking, physical activity, and for women, menopause status and hormone replacement treatment (HRT) (full model).

Stratified analyses were conducted according to the median age at infection (<70, ≥70 years), sex (male, female), BMI (<30, ≥30 kg/m^2^), physical activity (≤median, >median), and smoking status (never, ever) in the full models. To estimate potential effect modification by these stratification variables, we used a likelihood ratio test comparing the models with and without interaction terms between hormone levels and each stratification variable. In addition, sensitivity analysis was performed by excluding participants with self-rated poor overall health at baseline.

All analyses were performed with SAS 9.4. Statistical tests were all two-sided, and *P* <0.05 was regarded as statistically significant.

## Results

Among 678 male and 628 female patients with COVID-19, 194 (28.6%) and 121 (19.3%) deaths occurred, respectively. The median time from blood draw to COVID-19 testing was 11.1 years (interquartile range: 10.5-11.9 years). TT concentrations ranged from 1.96-27.45 nmol/L in men and 0.35-4.44 nmol/L in women (**Figure S1**). SHBG concentrations ranged from 2.59-133.66 nmol/L in men and 10.15-218.89 nmol/L in women.

**Table 1** summarizes the main characteristics of COVID-19 patients by quartiles of TT levels. FT and SHBG levels increased with TT levels. Moreover, patients with higher TT tended to be younger at blood draw. Men with higher BMI and women using HRT were more likely to have lower TT levels.

**Table 1.**
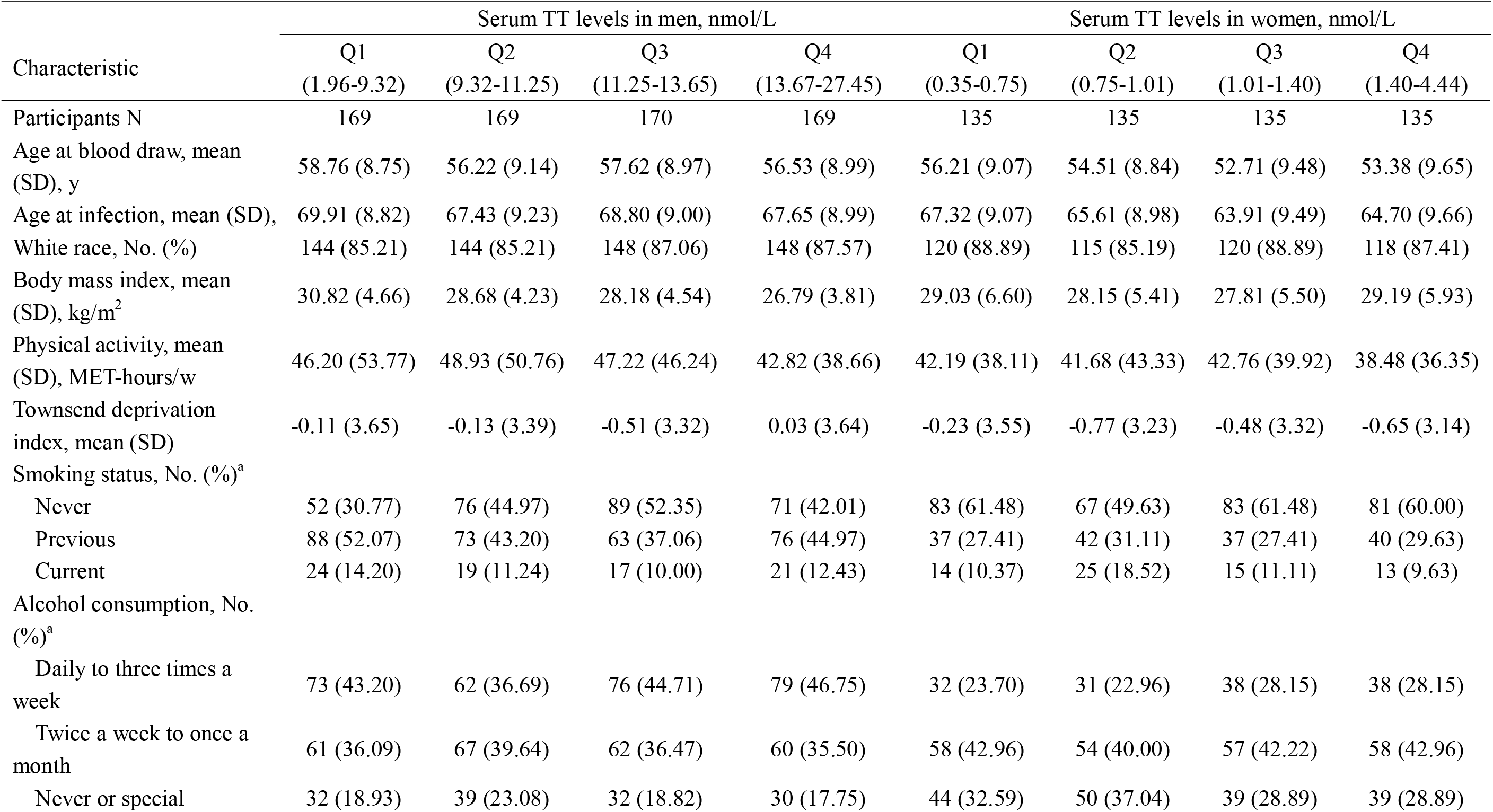
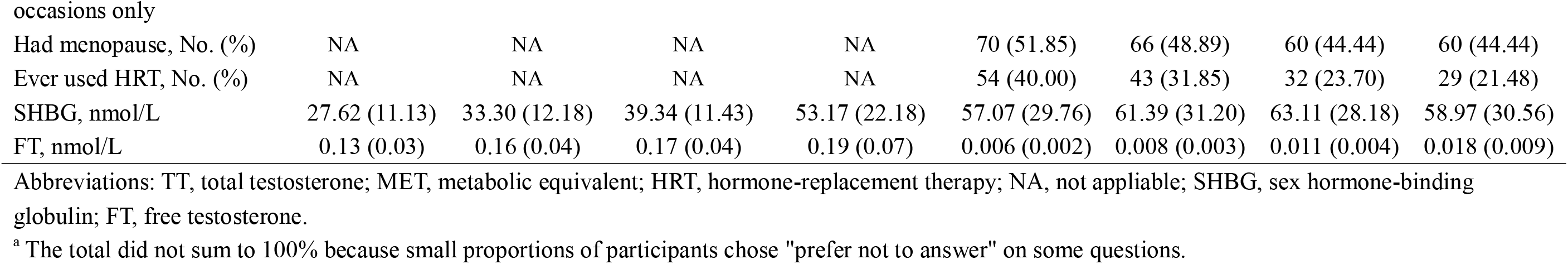
Baseline characteristics of COVID-19 patients according to quartiles of serum TT levels in men and women.

**Tables 2 and 3** showed the association results for the hormones and COVID-19 mortality in men and women, respectively. We did not find any statistically significant associations for TT (fully adjusted OR per 1-SD increase = 1.03, 95% CI: 0.85-1.25), FT (fully adjusted OR per 1-SD increase = 0.95, 95%CI: 0.77-1.17), or SHBG (fully adjusted OR per 1-SD increase = 1.09, 95% CI: 0.87-1.37) with COVID-19 mortality in men. Similar null results were observed in women (TT: fully adjusted OR per 1-SD increase = 1.10, 95% CI: 0.85-1.42; FT: fully adjusted OR per 1-SD increase = 1.10, 95% CI: 0.82-1.46; SHBG: fully adjusted OR per 1-SD increase = 1.16, 95% CI: 0.89-1.53).

**Table 2.**
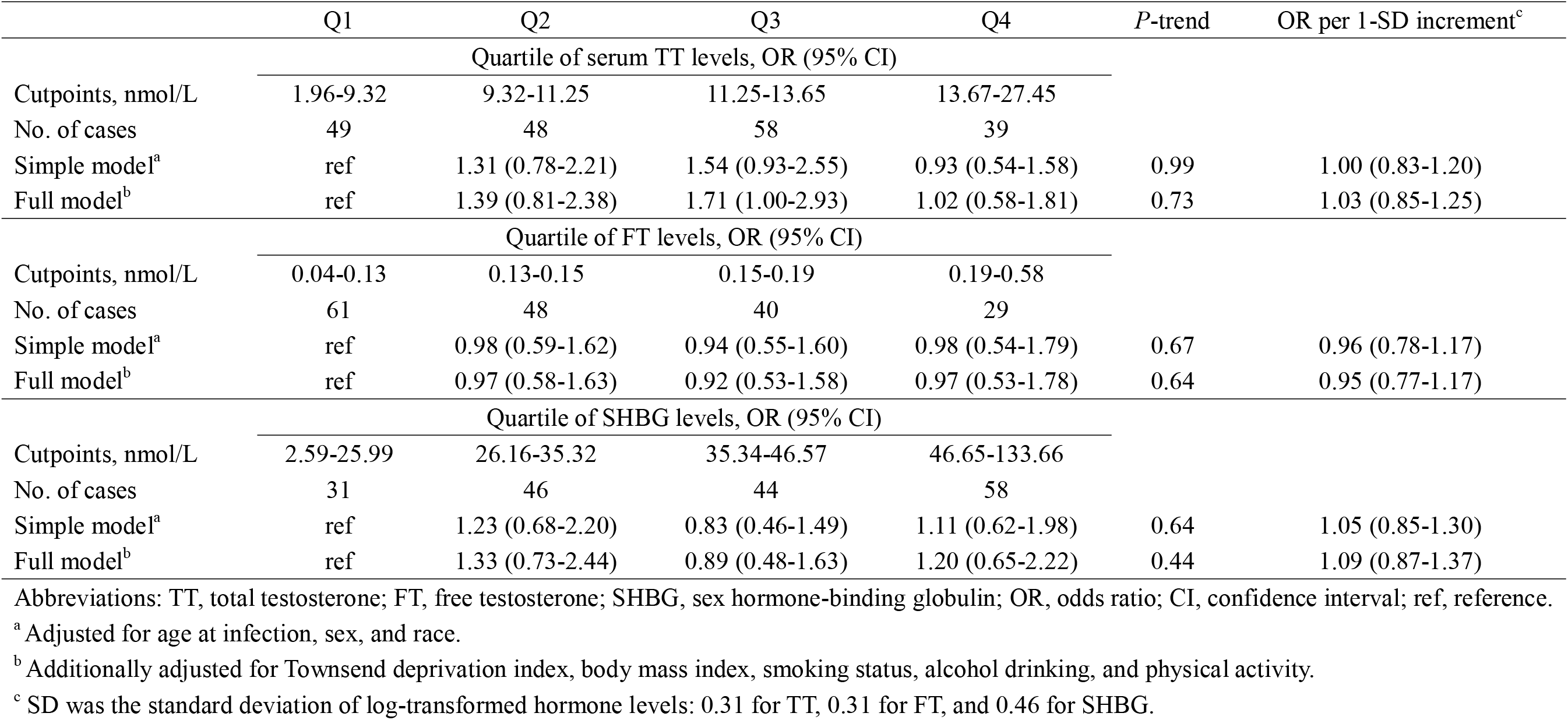
Associations of serum TT, FT, and SHBG levels with COVID-19 mortality in men.

|  | Q1 | Q2 | Q3 | Q4 | *P*-trend | OR per 1-SD increment<sup>c</sup> |
| --- | --- | --- | --- | --- | --- | --- |
| Quartile of serum TT levels, OR (95% CI) |  |  |  |  |  |  |
| Cutpoints, nmol/L | 1.96-9.32 | 9.32-11.25 | 11.25-13.65 | 13.67-27.45 |  |  |
| No. of cases | 49 | 48 | 58 | 39 |  |  |
| Simple model<sup>a</sup> | ref | 1.31 (0.78-2.21) | 1.54 (0.93-2.55) | 0.93 (0.54-1.58) | 0.99 | 1.00 (0.83-1.20) |
| Full model<sup>b</sup> | ref | 1.39 (0.81-2.38) | 1.71 (1.00-2.93) | 1.02 (0.58-1.81) | 0.73 | 1.03 (0.85-1.25) |
| Quartile of FT levels, OR (95% CI) |  |  |  |  |  |  |
| Cutpoints, nmol/L | 0.04-0.13 | 0.13-0.15 | 0.15-0.19 | 0.19-0.58 |  |  |
| No. of cases | 61 | 48 | 40 | 29 |  |  |
| Simple model<sup>a</sup> | ref | 0.98 (0.59-1.62) | 0.94 (0.55-1.60) | 0.98 (0.54-1.79) | 0.67 | 0.96 (0.78-1.17) |
| Full model<sup>b</sup> | ref | 0.97 (0.58-1.63) | 0.92 (0.53-1.58) | 0.97 (0.53-1.78) | 0.64 | 0.95 (0.77-1.17) |
| Quartile of SHBG levels, OR (95% CI) |  |  |  |  |  |  |
| Cutpoints, nmol/L | 2.59-25.99 | 26.16-35.32 | 35.34-46.57 | 46.65-133.66 |  |  |
| No. of cases | 31 | 46 | 44 | 58 |  |  |
| Simple model<sup>a</sup> | ref | 1.23 (0.68-2.20) | 0.83 (0.46-1.49) | 1.11 (0.62-1.98) | 0.64 | 1.05 (0.85-1.30) |
| Full model<sup>b</sup> | ref | 1.33 (0.73-2.44) | 0.89 (0.48-1.63) | 1.20 (0.65-2.22) | 0.44 | 1.09 (0.87-1.37) |

Abbreviations: TT, total testosterone; FT, free testosterone; SHBG, sex hormone-binding globulin; OR, odds ratio; CI, confidence interval; ref, reference.
<sup>a</sup> Adjusted for age at infection, sex, and race.
<sup>b</sup> Additionally adjusted for Townsend deprivation index, body mass index, smoking status, alcohol drinking, and physical activity.
<sup>c</sup> SD was the standard deviation of log-transformed hormone levels: 0.31 for TT, 0.31 for FT, and 0.46 for SHBG.

**Table 3.**
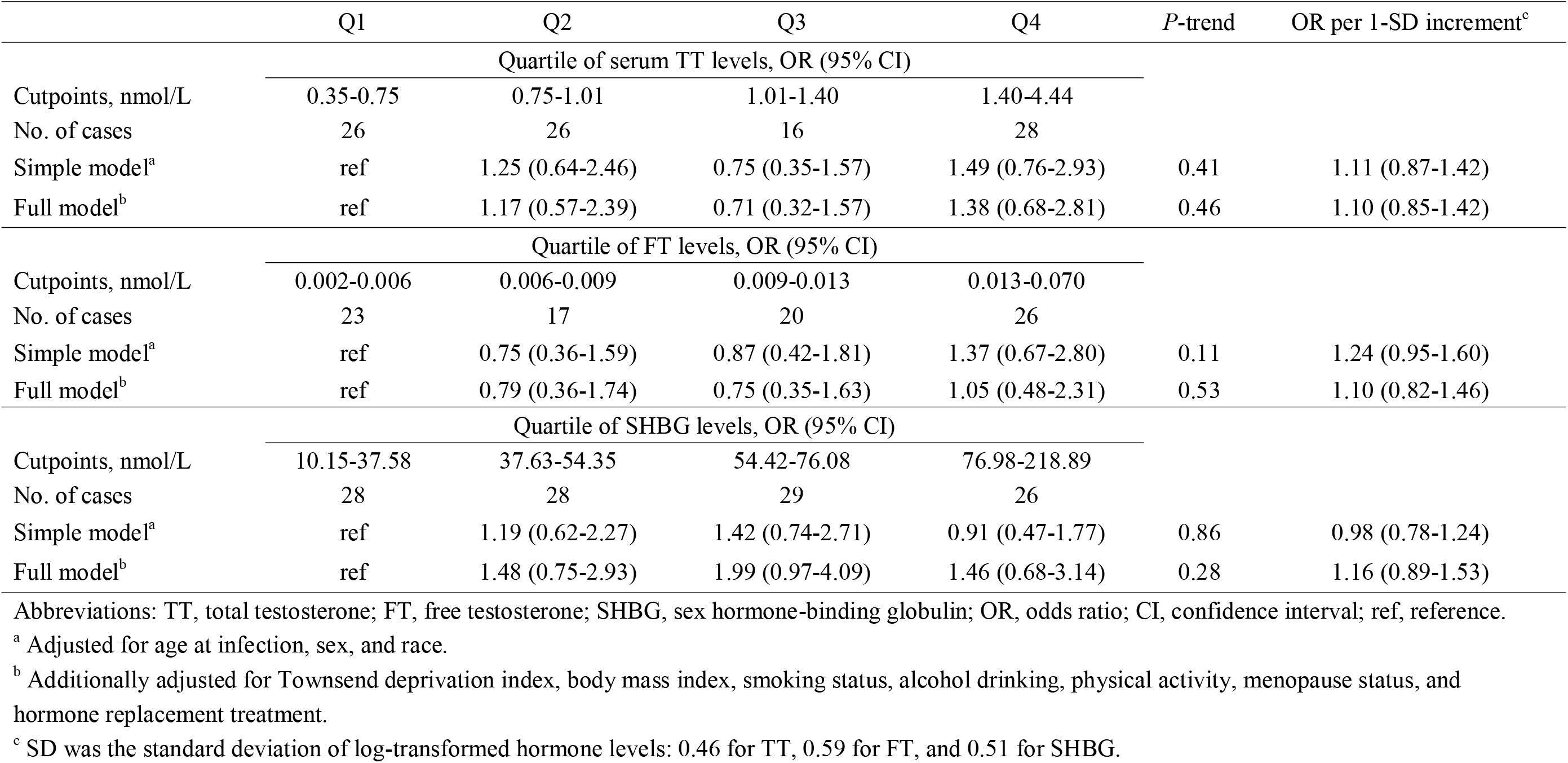
Associations of serum TT, FT, and SHBG levels with COVID-19 mortality in women.

|  | Q1 | Q2 | Q3 | Q4 | *P*-trend | OR per 1-SD increment<sup>c</sup> |
| --- | --- | --- | --- | --- | --- | --- |
| Quartile of serum TT levels, OR (95% CI) |  |  |  |  |  |  |
| Cutpoints, nmol/L | 0.35-0.75 | 0.75-1.01 | 1.01-1.40 | 1.40-4.44 |  |  |
| No. of cases | 26 | 26 | 16 | 28 |  |  |
| Simple model<sup>a</sup> | ref | 1.25 (0.64-2.46) | 0.75 (0.35-1.57) | 1.49 (0.76-2.93) | 0.41 | 1.11 (0.87-1.42) |
| Full model<sup>b</sup> | ref | 1.17 (0.57-2.39) | 0.71 (0.32-1.57) | 1.38 (0.68-2.81) | 0.46 | 1.10 (0.85-1.42) |
| Quartile of FT levels, OR (95% CI) |  |  |  |  |  |  |
| Cutpoints, nmol/L | 0.002-0.006 | 0.006-0.009 | 0.009-0.013 | 0.013-0.070 |  |  |
| No. of cases | 23 | 17 | 20 | 26 |  |  |
| Simple model<sup>a</sup> | ref | 0.75 (0.36-1.59) | 0.87 (0.42-1.81) | 1.37 (0.67-2.80) | 0.11 | 1.24 (0.95-1.60) |
| Full model<sup>b</sup> | ref | 0.79 (0.36-1.74) | 0.75 (0.35-1.63) | 1.05 (0.48-2.31) | 0.53 | 1.10 (0.82-1.46) |
| Quartile of SHBG levels, OR (95% CI) |  |  |  |  |  |  |
| Cutpoints, nmol/L | 10.15-37.58 | 37.63-54.35 | 54.42-76.08 | 76.98-218.89 |  |  |
| No. of cases | 28 | 28 | 29 | 26 |  |  |
| Simple model<sup>a</sup> | ref | 1.19 (0.62-2.27) | 1.42 (0.74-2.71) | 0.91 (0.47-1.77) | 0.86 | 0.98 (0.78-1.24) |
| Full model<sup>b</sup> | ref | 1.48 (0.75-2.93) | 1.99 (0.97-4.09) | 1.46 (0.68-3.14) | 0.28 | 1.16 (0.89-1.53) |

Abbreviations: TT, total testosterone; FT, free testosterone; SHBG, sex hormone-binding globulin; OR, odds ratio; CI, confidence interval; ref, reference.
<sup>a</sup> Adjusted for age at infection, sex, and race.
<sup>b</sup> Additionally adjusted for Townsend deprivation index, body mass index, smoking status, alcohol drinking, physical activity, menopause status, and hormone replacement treatment.
<sup>c</sup> SD was the standard deviation of log-transformed hormone levels: 0.46 for TT, 0.59 for FT, and 0.51 for SHBG.

The forest plot of stratified analyses (**Figure 1**) showed that none of the stratification variables had modification effects on TT, FT, and SHBG levels in men and women.

**Figure 1.**
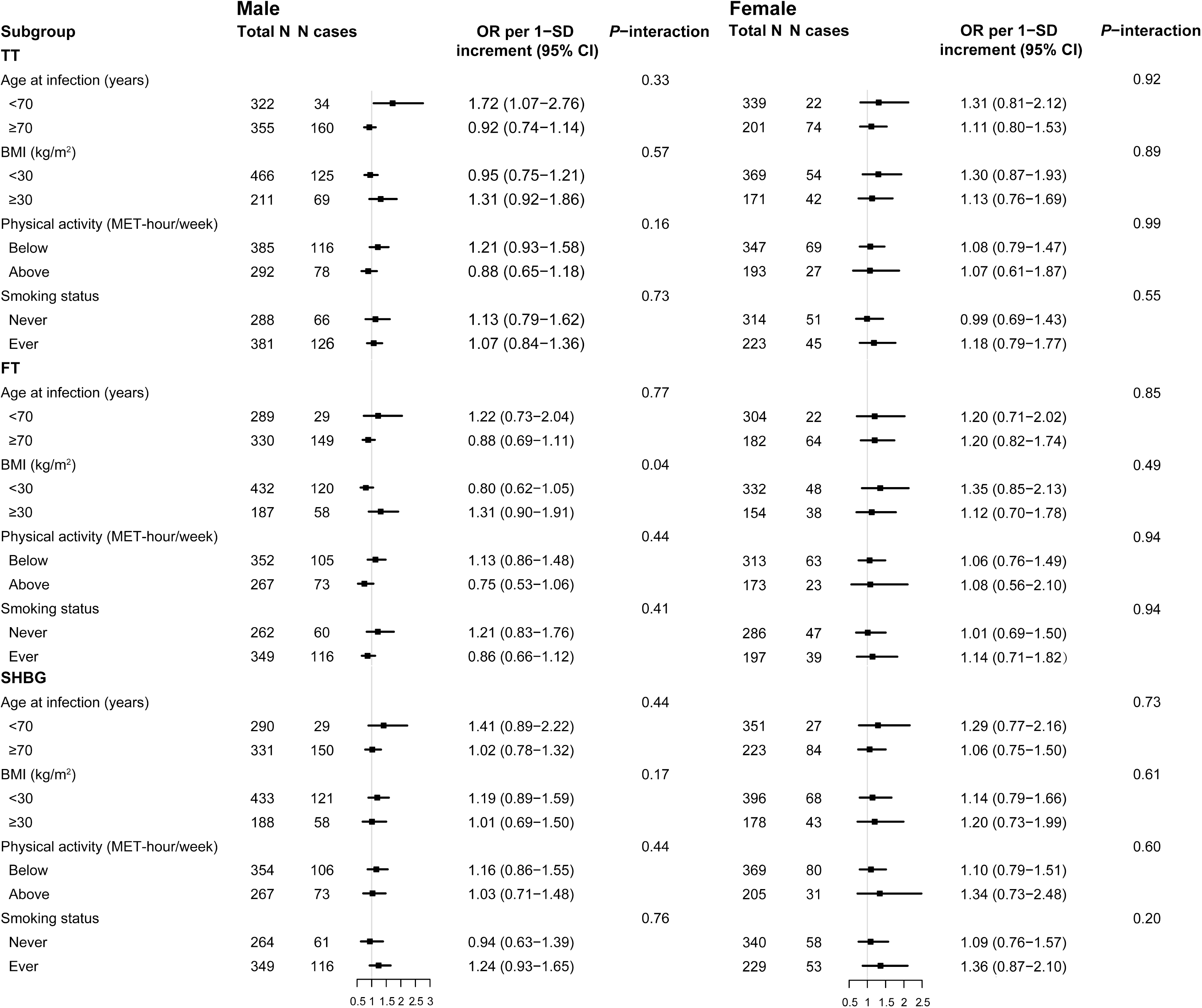
Forest plots of stratified analysis for the associations of serum total testosterone, free testosterone, and sex hormone-binding globulin levels with COVID-19 mortality.

The sensitivity analysis also indicated that the hormones were not associated with COVID-19 mortality (**Tables S1 and S2**).

## Discussion

To the best of our knowledge, this is the first study to assess the association of pre-diagnostic testosterone and SHBG levels with COVID-19 mortality. We did not observe any statistically significant associations in either men (n=678) or women (n=628), suggesting that circulating testosterone and SHBG may not play a critical role in COVID-19 prognosis.

In support of our findings, a prior study of 543 male patients with COVID-19 from UK Biobank did not identify any significant associations for baseline TT and SHBG levels with an increased risk of SARS-CoV-2 infection or hospitalization (12). However, in another study of 31 male patients with COVID-19 in Italy, lower TT and FT levels were associated with a higher risk of clinical deterioration (intensive care unit transfer or death) (13). Meanwhile, another study of 221 male patients reported that those who died from COVID-19 had lower mean TT levels than the patients alive (14). These two smaller studies were limited by inadequate adjustment for confounders, and they measured testosterone when patients were already hospitalized with COVID-19, which could introduce reverse causation.

The proposed leading cause of COVID-19 deterioration is the intense inflammatory response, known as a cytokine storm, which severely damages the lung tissue and causes acute respiratory distress syndrome (15). Sex differences in the response to inflammation have been well documented and can be attributed to various factors including sex hormones (16, 17). The current study suggests that circulating testosterone and SHBG may not be implicated in COVID-19 prognosis. However, we were unable to evaluate the association between estrogens and COVID-19 mortality because the majority of estradiol measurements were below the limit of detection in UK biobank. In a published cross-sectional study, serum estradiol showed negative correlations with severity of COVID-19 and levels of cytokines related to inflammation (18). Animal experiments also showed that estrogen signaling could suppress the inflammatory reactions and decrease severe acute respiratory syndrome coronavirus titers, leading to improved survival, while androgens did not influence disease outcome in male mice (19). Therefore, further studies are warranted to clarify whether and how estrogens have a beneficial effect on COVID-19 prognosis.

The major strengths of our study include the prospective design, reliable data on hormone levels and COVID-19, detailed data on covariates that allowed for robust confounding control, and evaluation of the associations both in men and women separately. However, two major limitations should be acknowledged. First, baseline hormones were assessed only one time, which may not reflect a long-time exposure. However, the intraclass correlation coefficients of TT and SHBG between two measurements (4 years apart in UK biobank) were 0.59 (95% CI: 0.58-0.61) and 0.86 (95% CI: 0.86-0.87), respectively, in men; 0.63 (95% CI: 0.62-0.64) and 0.81 (95% CI: 0.80-0.82), respectively, in women, indicating that a single measurement can reliably categorize average levels over at least a 4-year period (20). Second, limited by the coverage of COVID-19 testing, ascertainment bias may have occurred in the current study.

## Conclusions

Our findings indicate that circulating levels of testosterone and SHBG are not associated with COVID-19 prognosis. Future studies may focus on other factors to unreal the mechanisms underlying sex differences in COVID-19 mortality.

## Data Availability

The data that support the findings of this study are available from UK Biobank (https://www.ukbiobank.ac.uk/), but restrictions apply to their availability. These data were used under licence for the current study and so are not publicly available. The data are available from the authors upon reasonable request and with permission of UK Biobank.

http://biobank.ndph.ox.ac.uk/showcase/

## Acknowledgments

We are grateful to UK Biobank participants. This research has been conducted using the UK Biobank resource under application number 52217.

## Author Contributions

DH was responsible for the conception and design of the study. XF and DH had full access to all the data in the study and took responsibility for the integrity of the data and the accuracy of the data analysis. XF and JY did the statistical analysis and drafted the manuscript. JW, CY, MZ, HM, GJ, ZH, and HS critically revised the manuscript for important intellectual content. All authors reviewed and approved the final manuscript.

## Disclosure Summary

The authors have nothing to disclose.

**Abbreviations**
COVID-19: coronavirus disease 2019
SHBG: sex hormone-binding globulin
FT: free testosterone
TT: total testosterone
SARS-CoV-2: severe acute respiratory syndrome coronavirus 2
NHS: National Health Service
BMI: body mass index
MET: metabolic equivalent
ICD: international classification of diseases
HRT: hormone replacement treatment
OR: odds ratio
CI: confidence interval
SD: standard deviation.

